# Ruling In and Ruling Out COVID-19: Computing SARS-CoV-2 Infection Risk From Symptoms, Imaging and Test Data

**DOI:** 10.1101/2020.09.18.20197582

**Authors:** Christopher D’Ambrosia, Henrik I. Christensen, Eliah Aronoff-Spencer

## Abstract

**Background:** Assigning meaningful probabilities of SARS-CoV-2 infection risk presents a diagnostic challenge across the continuum of care.

**Methods:** We integrated patient symptom and test data using machine learning and Bayesian inference to quantify individual patient risk of SARS-CoV-2 infection. We trained models with 100,000 simulated patient profiles based on thirteen symptoms, estimated local prevalence, imaging, and molecular diagnostic performance from published reports. We tested these models with consecutive patients who presented with a COVID-19 compatible illness at the University of California San Diego Medical Center over 14 days starting in March 2020.

**Results:** We included 55 consecutive patients with fever (78%) or cough (77%) presenting for ambulatory (n=11) or hospital care (n=44). 51% (n=28) were female, 49% were age <60. Common comorbidities included diabetes (22%), hypertension (27%), cancer (16%) and cardiovascular disease (13%). 69% of these (n=38) were RT-PCR confirmed positive for SARS-CoV-2 infection, 11 had repeated negative nucleic acid testing and an alternate diagnosis. Bayesian inference network, distance metric-learning, and ensemble models discriminated between patients with SARS-CoV-2 infection and alternate diagnoses with sensitivities of 81.6 – 84.2%, specificities of 58.8 – 70.6%, and accuracies of 61.4 – 71.8%. After integrating imaging and laboratory test statistics with the predictions of the Bayesian inference network, changes in diagnostic uncertainty at each step in the simulated clinical evaluation process were highly sensitive to location, symptom, and diagnostic test choices.

**Conclusions:** Decision support models that incorporate symptoms and available test results can help providers diagnose SARS-CoV-2 infection in real world settings.

## INTRODUCTION

Despite advances in molecular diagnostics and imaging, ruling in or ruling out COVID-19 infection in an individual patient remains a significant challenge.^1^ Current CDC guidelines recommend providers determine whether signs or symptoms are compatible with COVID-19 infection and to test appropriate patients using nucleic acid amplification tests (NAATs) or antigen detection assays.^2^ However, diverse clinical presentations of COVID-19 infection may mimic those of common infections, potentially confounding the diagnostic value of presenting symptoms.^3^ Moreover, significant and evolving differences in estimated local disease prevalence for both COVID-19 infection and seasonal respiratory illnesses meaningfully impact differential diagnostic probabilities. Despite the uncertain utility of this symptom and local prevalence information, in low-resource and community settings such as ambulatory clinics, nursing homes, and telemedicine, these may be the only sources of data. In higher-resource settings, Nucleic Acid Amplification Tests (NAATs),^4^antibody based lateral flow assays,^5^chest radiography (CXR),^6^and computed tomography (CT)^7^may be available, yet published literature notes varied performance. Despite these limitations, clinicians with access to any imaging and testing modalities must optimize diagnostic imaging and testing sequences to appropriately reduce diagnostic uncertainty in a given setting.

This complexity highlights the need for reliable and user-friendly clinical decision support systems (CDSS) that suggest optimal testing strategies and quantify SARS-CoV-2 infection risk for patients across the continuum of care. Prior research has demonstrated the potential utility of Bayesian inference^8,9^and machine learning^10,11^methods in diagnostic decision-making, but computational clinical decision support has often been underutilized due to a lack of accessibility, transparency, workflow integration, and most importantly, the flexibility to incorporate local provider beliefs into the diagnostic model.^12,13^

A robust diagnostic risk model should be built on individualized patient data that is easily obtained by patients and healthcare workers. Menni et al. analyzed a large database of smartphone-enabled, self-reported symptom tracker records to predict potential COVID-19 cases using logistic regression models.^14^In the US test set, this approach had a reported sensitivity of 66% and a specificity of 83%. However, the performance of this or other machine learning models in clinical settings has not yet been examined.

Moreover, in evolving contexts where illness presentation may change depending on host and viral characteristics, large databases of individual patient records may not be available or locally-relevant. Constructing inflexible predictive algorithms, such as logistic regression models, based on out-of-date and locally irrelevant datasets would significantly compromise diagnostic accuracy. Addressing these issues, Chishti et al. demonstrated the advantages of using flexible probabilistic frameworks built without large-scale clinical datasets to generate ranked differential diagnoses that are more accurate that those developed by physicians.^15^

Combining the approaches of this prior work suggests that an appropriate diagnostic support model should rely on easily obtained symptom data, probabilistic frameworks to avoid the need for large-scale datasets, and most importantly, a flexible schema to refine predictions based on provider judgment and the ability to adapt to changes in local prevalence and current diagnostic test performance. To this end, we present a comparison and clinical validation of three such quantitative models as well as an ensemble approach to the diagnosis of COVID-19 in ambulatory and acute care settings. We then illustrate how this approach can be employed to help providers optimally reduce diagnostic uncertainty through appropriate diagnostic test choices and update predictions based on local clinical context and test results as that are obtained. Finally, we provide an interactive, on-line resource to assess COVID-19 infection probability based on user-defined parameters such as local-disease prevalence, imaging and testing performance (covid-calc.ucsd.edu).

## METHODS

### Data Acquisition

National and state-specific confirmed case figures as of 07/02/2020 for COVID-19 were acquired from the Center for Systems Science and Engineering at Johns Hopkins University.^16^During our model training, validation and testing process, we assumed a national SARS-CoV-2 infection prevalence of 11.1% based on total confirmed cases of 5,438,325 in the U.S. as of 08/17/2020,^16^a population estimate of 328,239,523,^17^and an estimated reporting rate of 14.9%.^18,19,20^Prevalence and conditional symptom probabilities for diseases in the differential diagnosis were collected from CDC and literature estimates (Table S1). COVID-19 symptom probabilities were developed primarily from a 393-person consecutive patient series^21^and supplemented by three meta-analyses which included 3,062,^22^49,504,^23^and 53,000 patients.^24^Where conditional symptom probabilities have not been described in the literature, we used a symptom probability of 1.0% based on our assumption that a higher conditional symptom probability would have been discussed in the literature.

To incorporate location and diagnostic test results into risk predictions, we used state-level case figures,^16^state-level population data,^17^and the estimated reporting rate^18,19,20^to compute an estimated SARS-CoV-2 infection prevalence for each state. We sourced imaging diagnostic accuracies from existing literature^6,7^and laboratory test accuracies from the Johns Hopkins Center for Health Security. RT-PCR sensitivity of 70% is based on published estimates^4^that take into account operator-dependency and variability in viral load across upper respiratory tract sites.^25^RT-PCR specificity of 99.8% is based on published data from Abbott Molecular.^26^Antibody test sensitivity and specificity based on published figures by Roche for electro-chemiluminescence immunoassay completed between 0-6 days of infection.^27^We computed likelihood ratios and prevalence-adjusted predictive values based on sensitivity, specificity and our estimated national COVID-19 prevalence of 11.1% (Table 1).

**Table 1.** Imaging and Laboratory Diagnostic Test Statistics for SARS-CoV-2 Infection (%).

|  | Sensitivity | Specificity | Likelihood Ratio |  | Predictive Value<sup>+</sup> |  |
| --- | --- | --- | --- | --- | --- | --- |
|  |  |  | Positive | Negative | Positive | Negative |
| **Chest Radiography** | 69.0<sup>Error!</sup><br>Bookmark not defined. | 50.0* | 1.4 | 0.6 | 14.7 | 92.8 |
| **Computed Tomography**<sup>Error! Bookmark not defined.</sup> | 97.0 | 57.3 | 2.3 | 0.1 | 22.1 | 99.3 |
| **RT-PCR** | 70.0<sup>Error!</sup><br>Bookmark not defined. | 97.0<sup>26</sup> | 23.3 | 0.3 | 74.5 | 96.3 |
| **Antibody**<sup>Error! Bookmark not defined.</sup><br>(0 – 6 days) | 65.5 | 99.8 | 327.5 | 0.3 | 97.6 | 95.9 |

\* No published figures available.
+ Prevalence-adjusted predictive values assume prevalence of 11.1%

### Training

We developed Bayesian inference network (BN) and set-cover (SC) models from published disease prevalence and conditional symptom probabilities (see supplemental methods S1). We simulated symptom profiles and diagnoses for 100,000 patients using the published aggregate diagnosis prevalence and conditional symptom probabilities. Of 100,000 simulated patients, the number of patients assigned to each mutually-exclusive diagnosis was proportional to diagnosis prevalence. Within each diagnostic class, we simulated a joint symptom distribution by randomly assigning the presence or absence of each symptom to every patient. The number of patients with a positive symptom within each diagnostic class was proportional to the conditional symptom probability for that symptom and diagnosis. We trained our DML and ensemble models on this simulated data.

### Study Design

We analyzed consecutive ambulatory and hospitalized patients with COVID-19 compatible syndromes presenting to UCSD Medical Center over 14 days in March and April 2020 with Institutional Review Board (IRB) approval (IRB # 200498). Patients were included if they had a recorded presenting illness including fever or cough, and at least a single NAAT in the electronic health record. Patients were labeled ‘positive’ if they had one or more positive RT-PCR tests and a compatible syndrome or findings on radiographic imaging. Patients were labeled ‘negative’ if they had 2 or more consecutive negative NAAT tests (>72 hours apart) or a single negative PCR and a negative antibody test within 14-21 days of symptom onset. Chart review was performed manually by an infectious disease specialist with an anonymized and blinded dataset presented for analysis (see Supplemental Methods for additional detail).

### Data Analysis

We calculated the sensitivity, specificity and prevalence-adjusted accuracy as well as the prevalence-adjusted negative predictive value (NPV) and positive predictive value (PPV) of each model on the clinical test data using standard Wald-type confidence intervals.^28^We estimated the 95% confidence intervals for sensitivity and specificity using Clopper-Pearson exact binomial proportion confidence intervals.^28^We estimated 95% confidence intervals for accuracy using the normal approximation method.^28^For the imaging and laboratory tests, we computed likelihood ratios based on sensitivity and specificity and prevalence-adjusted predictive values based on sensitivity, specificity, and an assumed national COVID-19 prevalence of 11.1%.

## RESULTS

### Patient Characteristics

Fifty-five individuals and the presence or absence of thirteen symptoms at initial presentation were included in our clinical test dataset. Thirty-eight patients (69.1%) were confirmed SARS-CoV-2 infection positive by RT-PCR. Forty-four subjects were seen on inpatient services, and 11 were seen as outpatients. 78.2% of subjects presented with fever, 63.6% with shortness of breath or dyspnea, 54.5% with non-productive cough, 21.8% with productive cough, 50.9% with fatigue or exhaustion, 9.1% with loss of smell, 7.3% with sore throat or pharyngalgia, 18.2% with body or muscle aches, 16.4% with headaches, 16.4% with diarrhea, 14.5% with nausea, 5.5% with vomiting, and 3.6% with nasal congestion or rhinorrhea. (Table 2, Table S2).

**Table 2.**
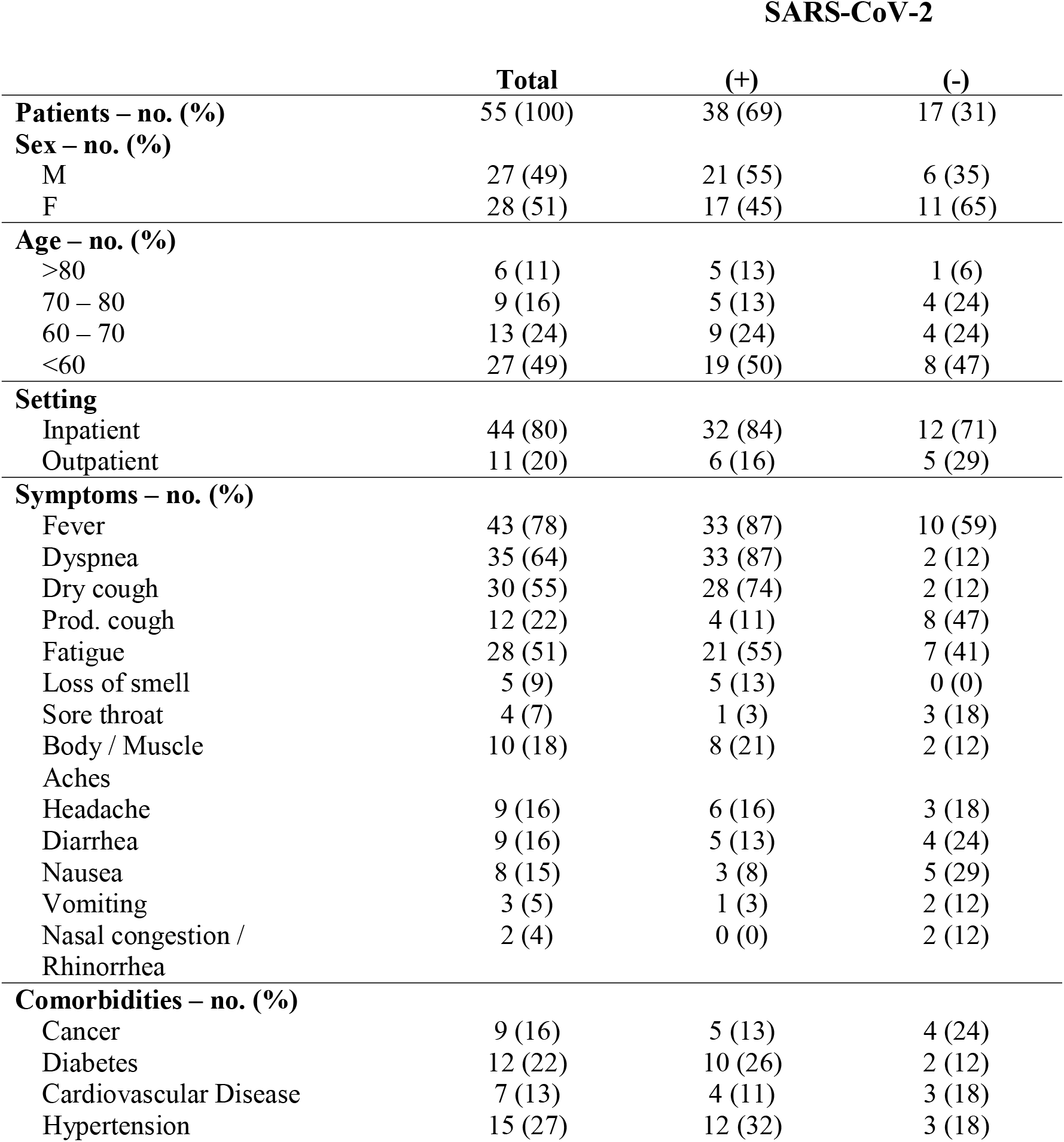
Clinical Test Dataset: Patient Characteristics.

### Classification Performance in the Clinical Test Dataset

Base models classified SARS-CoV-2 infection in the clinical test dataset with sensitivities and specificities of 81.6% (95% confidence interval [CI], 65.7 to 92.3) and 58.8% (95% CI, 32.9 to 81.6) for the BN model, 0.0% (95% CI, 0.0 to 9.3) and 100.0% (95% CI, 80.5 to 100.0) for the SC model, 84.2% (95% CI, 68.7 to 94.0) and 64.7% (95% CI, 38.3 to 85.8) for

the DML model, and 81.6% (95% confidence interval [CI], 65.7 to 92.3) and 70.6% (95% CI, 44.0 to 89.7) for the ensemble model. The overall accuracy of each of these models was 61.4% (95% CI, 48.5 to 74.2) for the BN model, 88.9% (95% CI, 80.6 to 97.2) for the SC model, 66.9% (95% CI, 54.4 to 79.3) for the DML model, and 71.8% (95% CI, 59.9 to 83.7) for the ensemble model. The prevalence-adjusted positive and negative predictive values for each model were 19.9% (95% CI, 10.5 to 29.2) and 96.2% (95% CI, 92.5 to 100.0) for the BN model, 0.0% and 88.9% for the SC model, 23.0% (95% CI, 11.3 to 34.7) and 97.0% (95% CI, 93.8 to 100.0) for the DML model, and 25.8% (95% CI, 11.4 to 40.2) and 96.8% (95% CI, 93.7 to 100.0) for the ensemble model (Table 3).

**Table 3.**
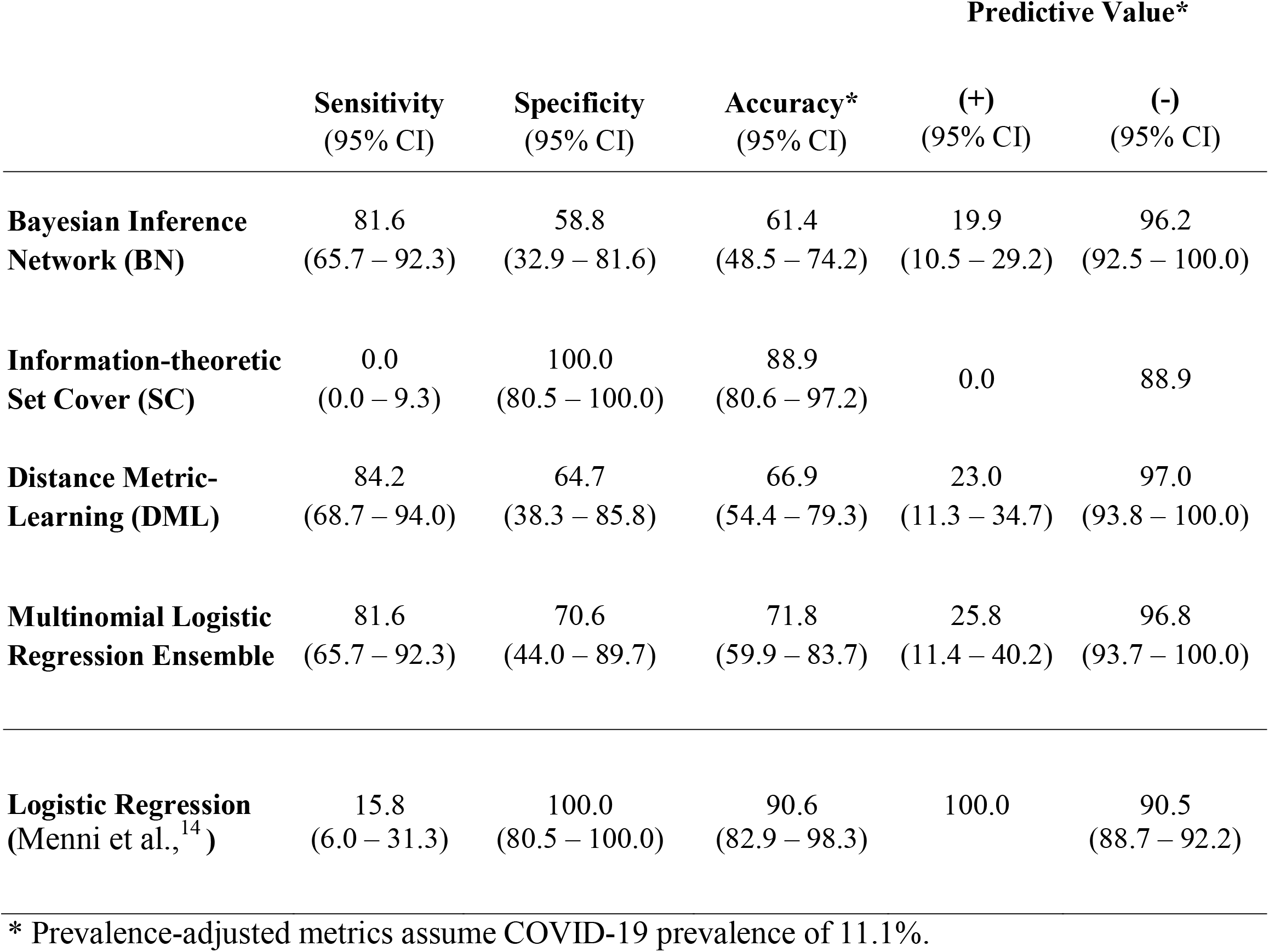
Classification performance on the Clinical Test Dataset for developed Base and Ensemble Models compared to a Logistic Regression Model reported in the literature (%).

### Incorporation of Location and Diagnostic Test Sequences

We then employed the BN model to evaluate three hypothetical patients with three different presentations: (1) fever, dry cough, shortness of breath, and anosmia; (2) fever and dry cough; (3) asymptomatic. We assumed all of these patients presented for care in area with a local disease prevalence equivalent to the national disease prevalence of 11.1%. For patient (1), we simulated a clinically plausible imaging and test result sequence of negative RT-PCR, negative antibody, and negative CXR. The probability of COVID-19 diagnosis following symptom collection was 99.8%. Despite negative test results, residual risk due to local disease prevalence and symptoms remained 97.7%. The change in diagnosis probability, or the reduction in diagnostic uncertainty, was only 2.1% following all three negative tests. For patient (2), we simulated the same negative test sequence. In this scenario, the combination of negative test results with non-specific symptom information resulted in a decrease in residual risk to 12.3%. The reduction in diagnostic uncertainty due to test results was 55.6%, primarily due to negative RT-PCR and negative antibody test results. The negative CXR provided less information as the reduction in diagnostic uncertainty following negative RT-PCR and antibody tests was only 6.2%. For patient (3), we simulated an imaging and test result sequence of negative RT-PCR, positive antibody, negative CXR. The negative RT-PCR test reduced disease probability by only 0.1%, and the positive antibody test increased the probability of COVID-19 diagnosis by 8.4%. The CXR results reduced diagnostic uncertainty by 3.0% (Figure 1).

**Figure 1.**
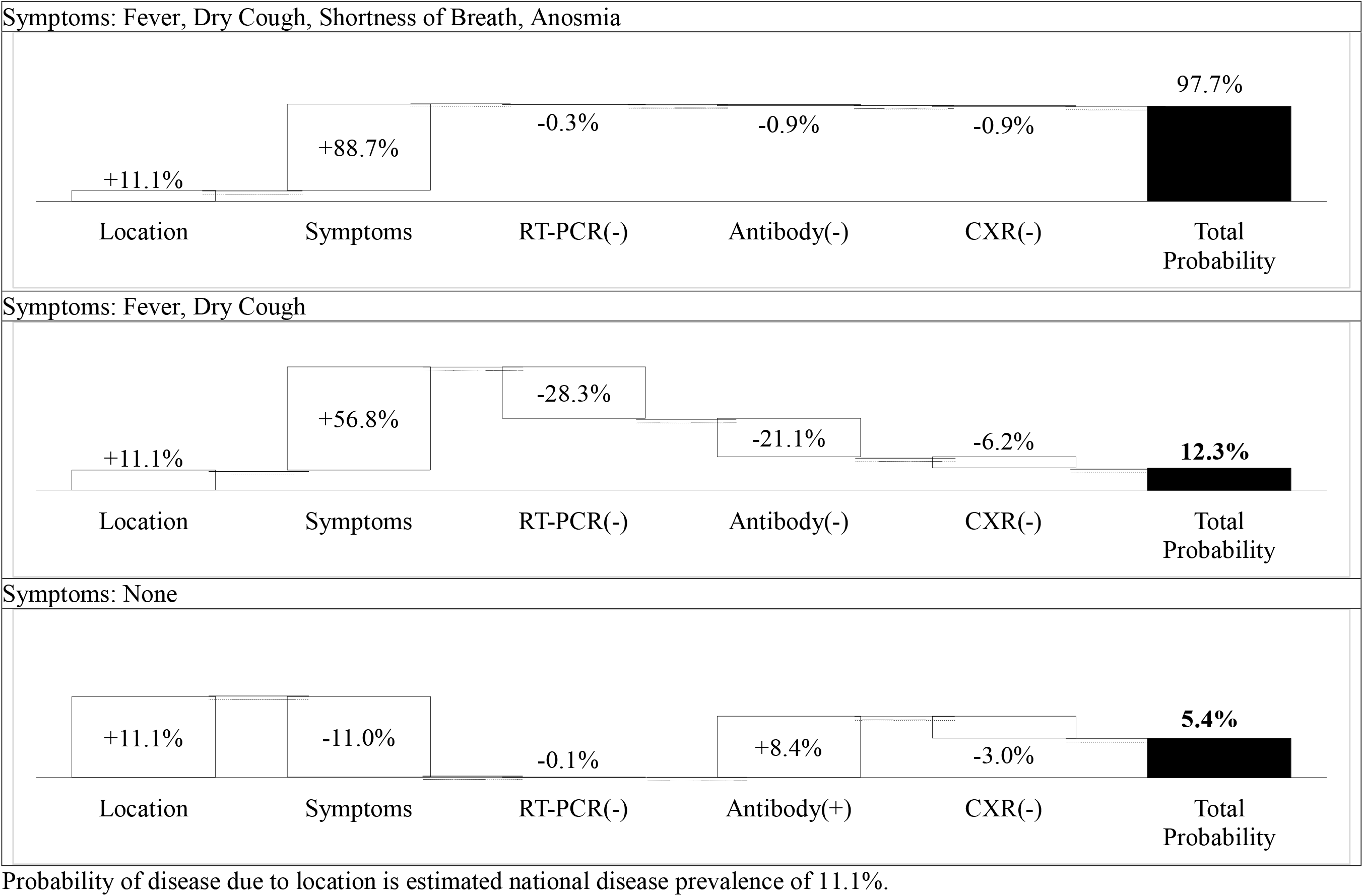
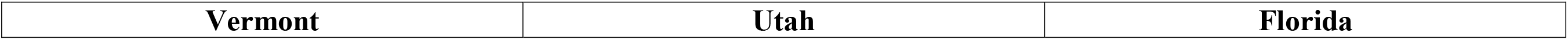
Probability of SARS-CoV-2 Infection for Common Patient Presentations and Clinical Test Sequences.

To illustrate the dependence of risk assessment on local disease prevalence, we simulated a patient with symptoms of only fever and dry cough presenting in three locations with significantly different COVID-19 prevalence estimates: Vermont (VT) with an estimated statewide prevalence of 1.6%, Utah (UT) with an estimated statewide prevalence of 9.8%, and Florida (FL) with an estimated statewide prevalence of 18.0% at the time of simulation. We combined results from three common test sequences with our BN pre-test probabilities to compute location-dependent risk trajectories. The test sequences included: (1) negative CXR and negative RT-PCR; (2) negative CXR and positive RT-PCR; (3) positive CXR and negative RT-PCR. Our results indicate that for a pauci-symptomatic patient presenting with identical symptoms in states with significantly different disease prevalence, post-test probabilities of SARS-CoV-2 infection following common diagnostic test sequences demonstrate marked variation. Moreover, changes in diagnostic probability or reductions in diagnostic uncertainty are highly-context and test dependent (Figure 2).

**Figure 2.**
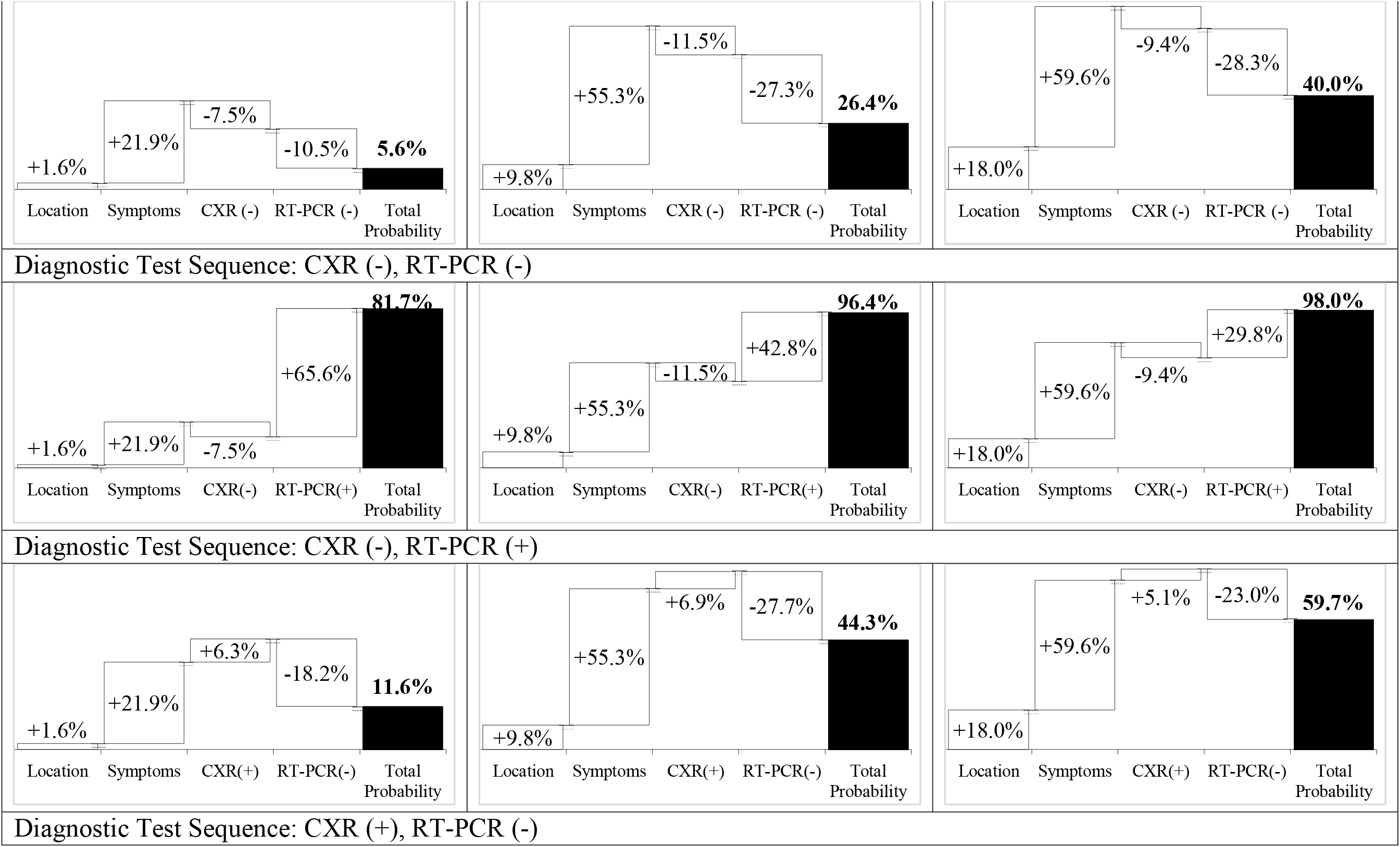
Impact of Patient Location and Diagnostic Test Results on Probability of SARS-CoV-2 Infection Probability of disease due to location is estimated disease prevalence for Vermont (1.6%), Utah (9.8%), and Florida (18.0%). Incremental probability due to symptoms assumes patient presents with only fever and dry cough.

## DISCUSSION

Our results suggest simple computable models that quantify patient risk of SARS-CoV-2 infection based on key elements of the clinical case can reduce diagnostic uncertainty for providers attempting to rule in or rule out disease with limited or conflicting information.

Building on work by Chishti et al.,^15^we chose probabilistic models in consideration of the scarcity of detailed, individual patient data and to take advantage of the depth of published literature on aggregate symptom probabilities. Our approaches to making step-wise diagnostic assessments with incremental information mimic clinical workflows and reflect the need for transparency and accommodation of new information critical to clinical decision-making. As in Menni et al.,^14^we chose clinical indicators that would be easily obtained by patients and providers as well as predictive models that are easily computed and transparent to all users. While other machine learning approaches, such as generative adversarial networks, transfer learning, n-shot learning, and prototypical networks, are also robust to limited data, these methods can be opaque and inaccessible to providers and may be inflexible and fragile in an evolving clinical context.

Our most simple model, the Bayesian inference network, is transparent, easily interpreted, and highly modifiable depending on the user’s prior beliefs about location-based prevalence, conditional symptom probabilities, and imaging and laboratory test accuracy. By developing base models that do not require access to large amounts of patient-level data and can accommodate changes in local provider beliefs and new sources of information, we alert physicians to the utility of using Bayesian reasoning to not only combine multiple data streams in order to make more informed diagnostic decisions, but also to guide decisions about use of imaging and testing that will most effectively reduce diagnostic uncertainty.

Our study has limitations. First, we used simulated patient data based on prevalence and conditional symptom probabilities to train and validate our DML and ensemble models that biased the ensemble model to heavily weight the DML model predictions. Second, the number of patients in our clinical test dataset was relatively small, and this dataset was enriched for SARS-CoV-2 positive patients due to the cancellation of all elective procedures and the use of telemedicine for almost all patient visits during the study period - leaving clinics and hospitals open primarily for COVID-19 patients and the acutely ill. Third, 80% of the patients in our clinical test dataset were from inpatient services, potentially biasing model accuracy by disease severity. Fourth, we chose as a reference standard the RT-PCR test results for SARS-CoV-2 infection despite outstanding questions about false negative rates in NAAT tests due to operator dependency and patient-level differences in viral loads across upper respiratory tract sites.^**Error! Bookmark not defined**.,25^

Overall, we found that Bayesian inference network, metric-learning model, and ensemble models trained and validated on a simulated patient dataset had sensitivities (81.6 – 84.2%) and specificities (58.8 – 70.6%) for discriminating between COVID-19 infection and other potential diagnoses in real clinical settings. These models had higher sensitivities than reported for most commonly used diagnostics, and model specificities were higher than those of both imaging modalities. For purposes of comparison, the logistic regression model proposed by Menni et al.,^14^when applied to our clinical test dataset, had a sensitivity of 15.8% and a specificity of 100.0%. Finally, the extension of our BN model shows that information acquired by imaging and testing choices is highly location and symptom dependent and emphasizes the utility of a quantitative framework to guide clinical decision-making in rapidly-changing local environments with potentially unreliable diagnostic tests.

## Data Availability

All data are available in the supplemental materials and methods.

https://covid-calc.herokuapp.com/

## ACKNOWLEDGEMENTS

We would like to thank Mary R. Mulcare, M.D at Weill Cornell School of Medicine, Dana R. Levin, M.D., M.P.H at Columbia University College of Physicians and Surgeons and Daniel L. Wang, M.D at the University of California San Diego School of Medicine, for scientific guidance, help with data preparation and manuscript comments.

## References

1. Song JY, Yun JG, Noh JY, et al. Covid-19 in South Korea - Challenges of subclinical manifestations. N Engl. J Med 2020;382(19):1858–9.

2. Coronavirus Disease 2019 (COVID-19) Testing Overview. Atlanta, GA: CDC, June 2020. (https://www.cdc.gov/coronavirus/2019-ncov/hcp/testing-overview.html)

3. Bordi L, Nicastri E, Scorzolini L, et al. Differential diagnosis of illness in patients under investigation for the novel coronavirus (SARS-CoV-2), Italy, February 2020. Eurosurveillance 2020;25(8):2000170.

4. Woloshin S, Patel N, Kesselheim AS. Perspective: false negative tests for SARS-CoV-2 infection - challenges and implications. N Engl J Med 2020. Epub June 5, 2020.

5. Global Progress on COVID-19 Serology-Based Testing. Baltimore, MD: Johns Hopkins Center for Health Security, June 2020. (https://www.centerforhealthsecurity.org/resources/COVID-19/serology/Serology-based-tests-for-COVID-19.html)

6. Wong HYF, Lam HYS, Fong AHT, et al. Frequency and Distribution of Chest Radiographic Findings in COVID-19 Positive Patients. Radiology 2019;201160.

7. Ai T, Yang Z, Hou H, et al. Correlation of Chest CT and RT-PCR Testing in Coronavirus Disease 2019 (COVID-19) in China: A Report of 1014 Cases. Radiology 2020;200642.

8. Grinfeld J, Nangalia J, Baxter EJ, et al. Classification and Personalized Prognosis in Myeloproliferative Neoplasms. N Engl J Med 2018;379(15):1416–30.

9. Palmerini T, Benedetto U, Bacchi-Reggiani L, et al. Mortality in patients treated with extended duration dual antiplatelet therapy after drug-eluting stent implantation: A pairwise and Bayesian network meta-analysis of randomised trials. Lancet 2015;385(9985):2371–82.

10. Milea D, Najjar RP, Jiang Z, et al. Artificial Intelligence to Detect Papilledema from Ocular Fundus Photographs. N Engl J Med 2020;382(18):1687–95.

11. Esteva A, Kuprel B, Novoa RA, et al. Dermatologist-level classification of skin cancer with deep neural networks. Nature 2017;542(7639):115–8.

12. Liu Y, Chen PHC, Krause J, Peng L. How to Read Articles That Use Machine Learning: Users’ Guides to the Medical Literature. JAMA 2019;322(18):1806–16.

13. Rosenbaum L, Lamas D. Cents and sensitivity. Teaching physicians to think about costs. N Engl J Med 2012;95(11):40–1.

14. Menni, C., Valdes, A.M., Freidin, M.B. et al.. Real-time tracking of self-reported symptoms to predict potential COVID-19. Nat Med 2020;26:1037–1040.

15. Chishti S, Jaggi KR, Saini A, Agarwal G, Ranjan A. Artificial Intelligence-Based Differential Diagnosis: Development and Validation of a Probabilistic Model to Address Lack of Large-Scale Clinical Datasets. J Med Internet Res 2020;22(4):e17550

16. Dong E, Du H, Gardner L. An interactive web-based dashboard to track COVID-19 in real time. Lancet Infect. Dis 2020;20(5):533–4.

17. State Population Totals and Components of Change: 2010-2019. Washington, DC: United States Census Bureau, December 2019. (https://www.census.gov/data/tables/time-series/demo/popest/2010s-state-total.html)

18. Commercial Laboratory Seroprevalence Data. Atlanta, GA: CDC, June 2020. (https://www.cdc.gov/coronavirus/2019-ncov/cases-updates/commercial-lab-surveys.html)

19. Havers FP, Reed C, Lim TW, et al. Seroprevalence of Antibodies to SARS-CoV-2 in Six Sites in the United States, March 23-May 3, 2020. medRxiv 2020;2020.06.25.20140384.

20. Pei S, Shaman J. Initial Simulation of SARS-CoV2 Spread and Intervention Effects in the Continental US. medRxiv 2020;2020.03.21.20040303.

21. Goyal P, Choi JJ, Pinheiro LC, et al. Clinical Characteristics of Covid-19 in New York City. N Engl J Med 2020;382(24):2372–4.

22. Zhu J, Ji P, Pang J, et al. Clinical characteristics of 3062 COVID-19 patients: A meta-analysis. J Med Virol 2020;jmv.25884.

23. Heydari K, Rismantab S, Shamshirian A, et al. Clinical and Paraclinical Characteristics of COVID-19 patients: A systematic review and meta-analysis. medRxiv 2020; 2020.03.26.20044057.

24. Ma C, Gu J, Hou P, et al. Incidence, clinical characteristics and prognostic factor of patients with COVID-19: a systematic review and meta-analysis. medRxiv 2020;2020.03.17.20037572.

25. Yu F, Yan L, Wang N, Yang S, et al. Quantitative Detection and Viral Load Analysis of SARS-CoV-2 in Infected Patients. Clin Infect Di. 2020. Epub Mar 28, 2020.

26. Current Molecular and Antigen Tests with FDA EUA Status. Baltimore, MD: Johns Hopkins Center for Health Security, June 2020. (https://www.centerforhealthsecurity.org/resources/COVID-19/molecular-based-tests/current-molecular-and-antigen-tests.html)

27. Global Progress on COVID-19 Serology-Based Testing. Baltimore, MD: Johns Hopkins Center for Health Security, June 2020. (https://www.centerforhealthsecurity.org/resources/COVID-19/serology/Serology-based-tests-for-COVID-19.html)

28. Mercaldo ND, Lau KF, Zhou XH. Confidence intervals for predictive values with an emphasis to case–control studies. Statistics in Medicine 2007;26(10):2170–83.

